# Mutation-specific impairment of TET2 and DNMT3A enzymatic activity predicts clonal hematopoiesis disease risk

**DOI:** 10.64898/2026.04.03.26350108

**Authors:** Yash Pershad, Kun Zhao, Joseph C Van Amburg, Robert W Corty, Alyssa C Parker, Alexander J Silver, Yara Almadani, Ashwin Kishtagari, Emily Hodges, Michael R Savona, J Brett Heimlich, Alexander G Bick

## Abstract

Clonal hematopoiesis of indeterminate potential (CHIP) driven by somatic mutations in TET2 and DNMT3A is present in >10% of adults over 60 and confers substantial risk for hematologic malignancy and cardiovascular disease, yet the majority of patients with CHIP do not progress to disease. Analyzing 1,020,538 individuals across three biobanks (UK Biobank, All of Us, BioVU), we show that a discrete subset of enzymatically disruptive mutations — *TET2* loss-of-function variants and the *DNMT3A* R882 hotspot — account for the majority of clinical risk in these genes and exhibit the strongest clonal fitness advantage. Because *DNMT3A* and *TET2* encode enzymes that modulate DNA methylation, we reasoned that peripheral blood methylation patterns should reflect the functional impact of individual mutations, enabling a direct readout of enzymatic dysfunction in CHIP patients. We developed and validated methylation-based activity scores for *TET2* and *DNMT3A* as patient specific biomarkers that quantify enzymatic activity. These scores capture functional heterogeneity across mutation subtypes, predict disease risk comparably to clinical risk scores such as the Clonal Hematopoiesis Risk Score and the AHA PREVENT cardiovascular risk model. Integrating the activity score with the clinical models substantially improves prediction of incident cytopenia, myeloid neoplasm, and major adverse cardiovascular events. These findings establish that *TET2* and *DNMT3A* CHIP pathogenicity is proportional to the degree of enzymatic disruption conferred by specific variants, and nominate methylation-based activity scores as a functional biomarker for individualized CHIP risk stratification and monitoring therapeutic response.

## Main

In clonal hematopoiesis of indeterminate potential (CHIP), hematopoietic stem cells acquire somatic mutations in driver genes, producing clones of peripheral blood cells^1,2^. CHIP is present in >10% of adults over 60 years of age and increases risk of cytopenia and myeloid neoplasm by 7-fold and atherosclerotic cardiovascular disease (ASCVD) by >1.5-fold, along with multiple other diseases associated with aging^2–5^.

It remains unclear why only a subset of individuals with CHIP progress to disease. Identifying patients at highest risk remains a critical barrier to clinical development and care delivery for those with CHIP^6^. Current predictive models for myeloid neoplasm risk, such as the Clonal Hematopoiesis Risk Score (CHRS)^7^ and MNPredict^8^ leverage demographic, clinical, and genetic factors, but treat all mutations within the same gene equally. On the other hand, risk stratification tools for atherosclerotic cardiovascular disease (ASCVD) which estimate 10-year risk, such as PREVENT and the Pooled Cohort Equation, do not include CHIP as a risk factor.^9,10^

While somatic mutations in at least 74 genes can cause CHIP, approximately 75% of them occur in one of just two genes, both of which encode enzymes that alter DNA methylation: DNA methyltransferase 3 alpha (*DNMT3A*) and Tet Methylcytosine Dioxygenase 2 (*TET2*)^1,3,5^. *DNMT3A* encodes a de novo DNA methyltransferase, which oligomerizes to establish de novo methylation marks at CpG dinucleotides, while *TET2* encodes an enzyme which catalyzes the oxidation of 5-methylcytosine to 5-hydroxymethylcytosine^11,12^. Pathogenic mutations in either gene are thought to disrupt the balance of methylation and demethylation, leading to epigenetic reprogramming in hematopoietic stem cells^13–16^.

Pathogenic mutations in *DNMT3A* and *TET2* compromise a mixture of missense, splicing, frameshift and stop-gain mutations^17^. Prior studies have highlighted significant heterogeneity of these mutations on protein stability and function^16^. The most well-characterized specific mutation is the *DNMT3A* R882 hotspot. The R882H mutation has been shown to exhibit 80% reduced methyltransferase activity and disrupt DNMT3A oligomerization in vitro^18^. R882 resides at a critical position within the DNMT3A protein where it is spatially arranged in the DNMT3A-DNMT3A homodimeric interface while additionally participating in DNA binding both directly through contacts with the DNA backbone as well as indirectly through stabilizing contacts with the protein’s target recognition domain loop^19,20^. In *TET2*, mutations in the catalytic domain are associated with increased methylation at key regulatory enhancer sites and global loss of hydroxymethylation^21–24^. However, since each mutation type within a driver gene is individually rare, prior work has lacked sufficient power to understand the mutation-specific clinical consequences within *DNMT3A* and *TET2*.

To address this, we analyzed >1 million individuals across three biobanks to characterize the mutational landscape, cellular fitness effects and clinical consequences of *TET2* and *DNMT3A* CHIP. We found that loss-of-function mutations in *TET2* and R882 mutations in *DNMT3A* confer significantly higher cellular fitness and disease risk across a broad spectrum of CHIP outcomes, implying that increased enzymatic disruption is linked to increased disease risk. Since *DNMT3A* and *TET2* CHIP patients have distinct DNA methylation signatures in peripheral blood^13^, we hypothesized that the methylation profile of peripheral blood could represent a functional readout of DNMT3A and TET2 activity and serve as a biomarker for relative mutant clone impact and disease risk. Leveraging both in vitro models and patient peripheral blood methylation profiles, we derived and validated enzymatic activity scores for TET2 and DNMT3A. Incorporating this individual level quantification of DNMT3A or TET2 enzymatic activity significantly enhances risk prediction across multiple diseases. Together these observations establish that the pathogenicity of *TET2* and *DNMT3A* CHIP is linked to the degree of underlying enzymatic disruption caused by specific variants, thereby nominating enzymatic activity as a clinically useful biomarker to quantify an individual’s CHIP-associated disease risk.

## Results

### Mutational landscape of TET2 and DNMT3A in CHIP differs from that in myeloid neoplasm

We analyzed 1,020,538 individuals with whole-genome or whole-exome sequencing from UK Biobank (UKB), NIH All of Us, and Vanderbilt BioVU and detected somatic mutations using Mutect2, filtering variants as described by Vlasschaert et al^5^ (**Extended Data Figure 1**). Among 846,353 individuals ≥40 years old, we identified 35,270 with CHIP (4.2%); these included 19,368 (2.3%) with *DNMT3A* mutations, and 6,762 (0.80%) with *TET2* mutations. By definition, these CHIP variants exhibited a strong association with age.

We compared the mutational landscape of *TET2* and *DNMT3A* in CHIP and 16,264 cases of myeloid neoplasm (MN) in the cBioPortal for Cancer Genomics (http://cbioportal.org)^25^. *TET2* mutations in CHIP and MN distribute across the protein without significant hotspots (**Figure 1A**), consistent with prior work^1,3,5^. Three amino acid residues in *TET2* were more frequently mutated in MN than CHIP: an arginine residue at position 1261 which participates in the binding of 2-oxoglutarate, and two residues – a cysteine at position 1358 and a histidine residue at position 1380 – which are part of the binding site of the second Zn^2+^ cofactor, which stabilizes TET2’s catalytic domain (**Figure 1B**; **Supplemental Table 1**)^26^. The glutamine residue at position 1368 and arginine residue at position 1394 were more frequently mutated in CHIP than MN. In aggregate, putative loss-of-function (pLoF) mutations were more common in MN compared to CHIP (74.0% vs 62.4%, P = 7×10^−34^) (**Figure 1C**).

**Figure 1:**
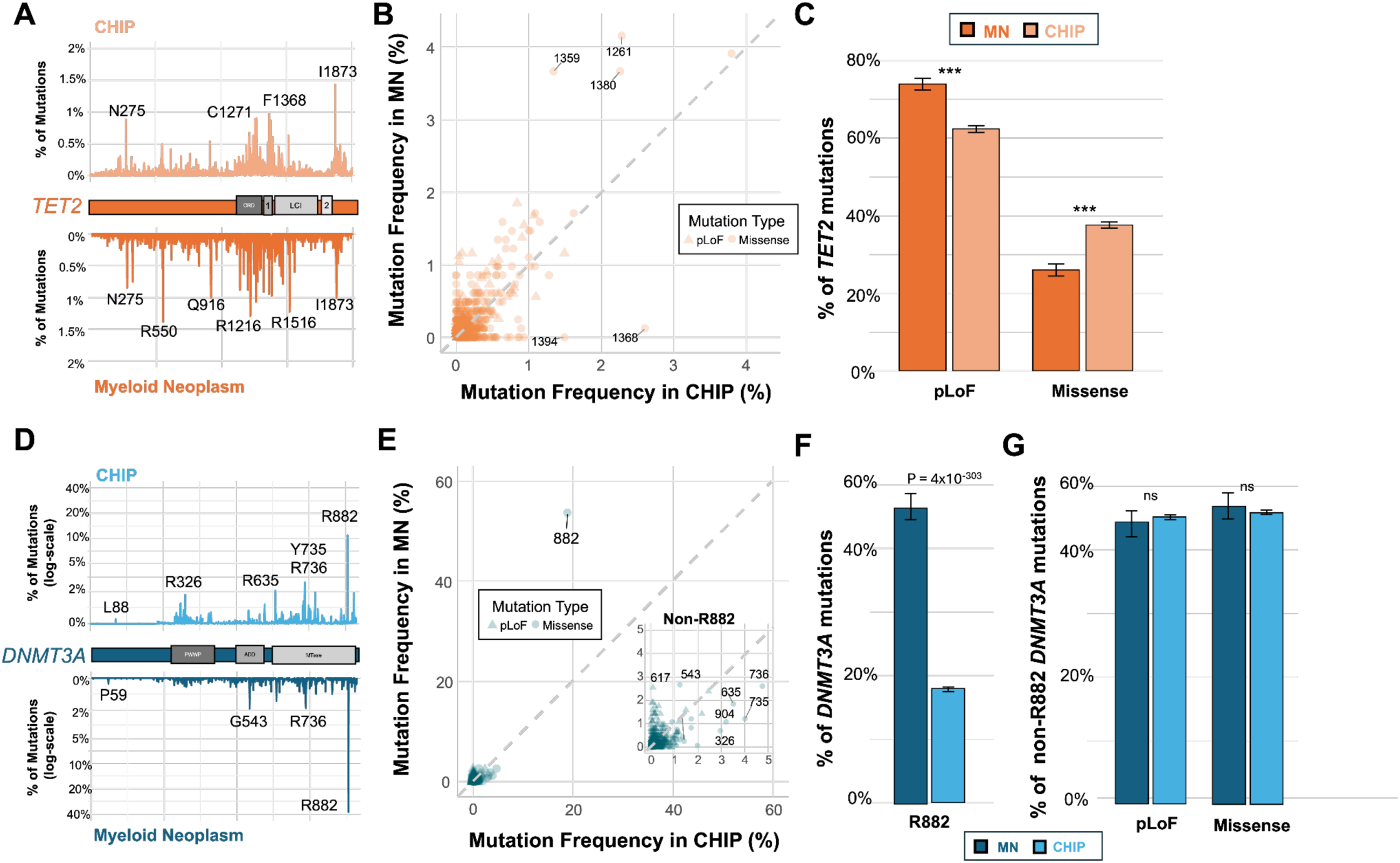
Comparative mutational landscape of TET2 and DNMT3A in clonal hematopoiesis (CHIP) and myeloid neoplasm (MN). a) Distribution of *TET2* somatic mutations across protein domains in CHIP (N=6,762) and MN (N=16,264). b) Residue-level enrichment analysis identifying specific amino acids in *TET2* (e.g., C1358, H1380) significantly more frequent in MN compared to CHIP. c) Percentage of *TET2* mutation types; protein-loss-of-function (pLoF) variants are significantly enriched in MN (74.0%) versus CHIP (62.4%; P=7×10^−34^ by two-sided Chi-squared test). d) Distribution of *DNMT3A* mutations, highlighting the dominance of the R882 hotspot in both cohorts. e) Comparison of residue frequency in *DNMT3A*; R882 mutations are significantly overrepresented in MN (53.7%) compared to CHIP (18.9%; P=4×10^−303^). f) Frequency of R882 mutations as a proportion of total *DNMT3A* variants. g) Relative proportions of pLoF and missense variants in *DNMT3A* following the exclusion of R882 mutations, demonstrating no significant (ns) difference between CHIP and MN cohorts. Data are presented as mean ± standard error.

*DNMT3A* mutations cluster predominantly at the well-known arginine hotspot at position 882 (R882) (**Figure 1D**). *DNMT3A* R882 mutations were more frequent in MN than in CHIP (53.7% vs 18.9%, P = 4×10^−303^) (**Figure 1E**; **Supplemental Table 2**; **Figure 1F**). After excluding R882, missense mutations at residue 543 and pLoF mutations at residue 617 were each significantly more frequent in MN than CHIP (6.7% versus 2.5%, P = 1.8×10^−9^ and 2.5% versus 0.1%, P < 10^−16^, respectively). There was no significant difference in the proportion of pLoF and missense mutations in *DNMT3A* in CHIP versus MN after excluding R882 mutations (**Figure 1G**).

### Only certain TET2 and DNMT3A mutations exhibit cellular drive for loss of heterozygosity

We next hypothesized that specific *TET2* and *DNMT3A* mutations had differential effects on cellular fitness, which we quantified based on the selection pressure for a mutation to become homozygous via a copy-neutral loss of heterozygosity (CN-LOH) of the region harboring the mutation (**Extended Data Figure 2**). Since the risk of CN-LOH of the mutation represents the strength of selection for the mutation at a cellular level, we would anticipate that mutations that more frequently appear in regions of CN-LOH confer a cellular fitness advantage.

Prior studies have shown that both *TET2* and *DNMT3A* increase risk of CN-LOH of their chromosomal arms, chr4q and chr2p respectively^27–29^. Consistent with prior work, both *DNMT3A* and *TET2* CHIP, when aggregating all mutations in the genes, significantly increased risk of CN-LOH mosaic chromosomal alterations on the same chromosomal arm. *TET2* conferred higher risk of CN-LOH than *DNMT3A*. Of the 6,762 people with *TET2* CHIP, 48 had detectable chr4q CN-LOH (odds ratio (OR): 20.0, 95% confidence interval (CI): 13.8 - 28.4, P= 3×10^−60^). Of the 19,368 people with *DNMT3A* CHIP, only 11 had detectable chr2p CN-LOH (odds ratio (OR): 4.13, 95% confidence interval (CI): 2.05-7.54, P= 2×10^−5^).

Prior studies to-date have not had sufficient power to evaluate CN-LOH co-occurrence with mutation-type resolution. Stratification by mutation type revealed that *TET2* protein-loss-of-function (pLoF) variants were more strongly associated with CN-LOH in magnitude than missense mutations (pLoF, OR: 19.72, 95% CI: 13.18-28.84, P=1.0×10^−50^, N=36; missense, OR: 8.7, 95% CI: 4.08-16.3, P=6.0×10^−10^, N=10) (**Extended Data Figure 2**; **Supplemental Table 3**). However, the rarity of CN-LOH events limited the statistical power to formally distinguish between these risk profiles. The number of CN-LOH events for individuals with both TET2 pLoF and missense mutations exceeded expectations (40 observed events versus 2 expected by chance, P = 4×10^−49^; 10 observed events versus 1 expected by chance, P = 1×10^−7^) (**Extended Data Figure 2**).

For *DNMT3A*, pLoF mutations had a higher than expected number of observed chr2p CNLOH events (3 observed versus 1 expected, P = 0.08) and an odds ratio of CN-LOH above 1, yet was not statistically significant (OR: 2.14, 95% CI: 0.52- 5.76, P = 0.20) (**Extended Data Figure 2**; **Supplemental Table 4**). Among missense mutations, only non-R882 missense mutations were significantly associated with increased risk of chr2p CNLOH (OR: 6.55, 95% CI: 2.40-10.7, P = 7×10^−6^). Strikingly, despite these being the most common missense mutations, *DNMT3A* R882 mutations never co-occurred with chr2p CN-LOH events (**Extended Data Figure 2**). These findings suggest *DNMT3A* R882 mutations lack a selection pressure towards homozygosity. This result is consistent with the hypothesized dominant negative function of R882 mutations^18^, as if R882-mutant DNMT3A disrupt wild-type DNMT3A enzymes when they oligomerize, the cell functionally already has nearly a complete lack of DNMT3A enzymatic activity.

### Clinical consequences of TET2 and DNMT3A CHIP are mutation-type-specific

We next hypothesized that disease risk for *TET2* and *DNMT3A* CHIP would vary by mutation type. To test this hypothesis, we performed a time-to-event analysis with Cox proportional hazards regression across the entire medical “phenome” across UKB, AllofUs, and BioVU by *TET2* and *DNMT3A* CHIP by mutation type.

For *TET2*, we observed that pLoF mutations were associated with increased disease risk, while missense variants showed no significant associations (**Supplemental Table 5**). To compare the effect sizes of TET2 pLoF and missense mutations across the phenome, we performed a mixed-effects meta-regression on 2,180 phenotypes. We observed that pLoF mutations exhibited significantly higher hazard ratios than missense mutations (β_diff_ = 0.06, SE = 0.008, P = 5×10^−15^), accounting for approximately 9.86% of the observed heterogeneity in effect sizes. Among 48 phenotypes significantly associated with *TET2* CHIP or its mutation-specific subtypes after meta-analysis with multiple-hypothesis correction, pLoF mutations conferred higher hazard ratios than missense mutations for 43 phenotypes (89.6%; one-sided sign test P = 7×10^−9^) . Among known phenotypic associations of *TET2* CHIP, including myeloid neoplasm^1,30^, bacterial infection^31^, venous thromboembolism (VTE)^32,33^, anemia^4^, acute kidney injury (AKI)^34^, chronic kidney disease (CKD)^35,36^, lymphoid neoplasm, and atherosclerotic cardiovascular disease (ASCVD)^2,37–39^, the hazard ratios (HRs) of pLoF mutations were higher than missense mutations for each (**Figure 2A**). We observed the same trend for incident persistent cytopenia^4^ (pLoF, HR: 1.58, 95% CI: 1.38-1.82, P = 7×10^−11^ versus missense, HR: 1.24, 95% CI: 0.76-2.03, P=0.38), and the five-point composite of major adverse cardiovascular events (MACE) (pLoF, HR: 1.40, 95% CI: 1.28-1.54, P = 3×10^−12^ versus missense, HR: 1.14, 95% CI: 0.97-1.33, P=0.10) (**Figure 2B**). Missense mutations confer substantially lower risk than pLoF, and, in many cases, do not reach the threshold for significant clinical association. These results demonstrate that the pathogenic risk of *TET2* CHIP is primarily concentrated in pLoF variants, with missense variants in aggregate exhibiting consistently attenuated effect sizes across a broad spectrum of clinical outcomes.

**Figure 2:**
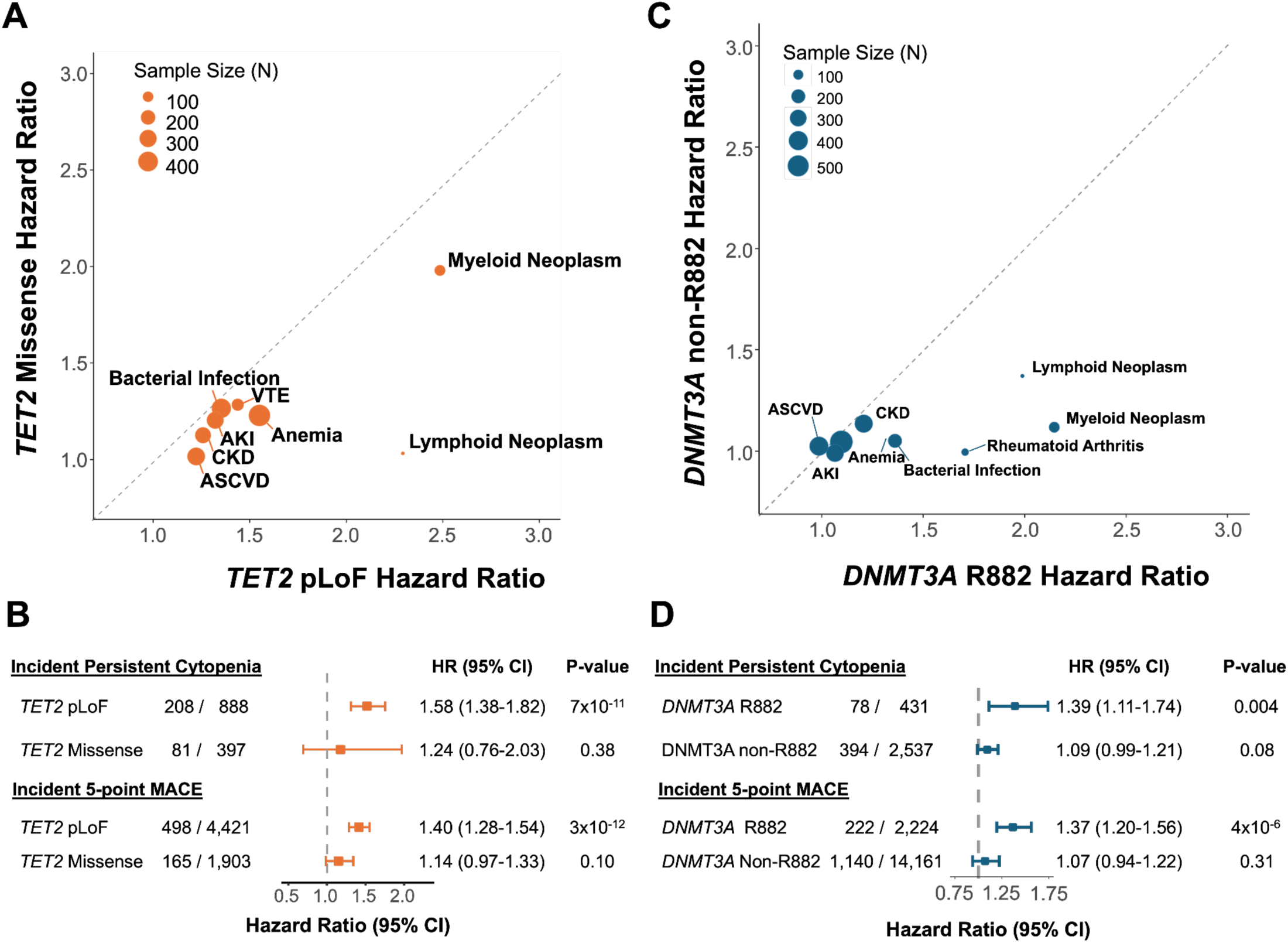
Variant-specific clinical risk profiles across the medical phenome. a) Meta-regression comparing hazard ratios (HR) for *TET2* pLoF versus missense mutations across diverse clinical outcomes. pLoF variants consistently confer higher risk for conditions including myeloid neoplasm, ASCVD, and VTE. b) Forest plots for incident persistent cytopenia and 5-point major adverse cardiovascular events (MACE) by *TET2* mutation type. c) Comparison of HRs for *DNMT3A* R882 versus non-R882 variants; risk is predominantly concentrated in the R882 hotspot. d) Impact of *DNMT3A* mutational archetype on cytopenia and MACE risk. Only R882 mutations significantly associate with increased incidence of these outcomes. P-values and HRs were derived from adjusted Cox proportional hazards models.

For *DNMT3A*, R882 mutations confer most of the disease risk, while other *DNMT3A* mutations were not associated with disease risk. We compared the phenotypic impact of the DNMT3A R882 hotspot mutation against other missense mutations across 2,180 phenotypes using a mixed-effects meta-regression. The R882 variant exhibited a significantly higher mean effect size (β_diff_ = 0.10, SE = 0.02, P = 1.35×10^−5^). Among 8 phenotypes significantly associated with *DNMT3A* CHIP or its mutation-specific subtypes in our analysis, R882 mutations conferred higher hazard ratios than non-R882 mutations for all 8 phenotypes (100%; one-sided sign test P=4×10^−3^) (**Supplemental Table 6**). Focusing on same phenotypes listed above, with the addition of rheumatoid arthritis which is known to be associated with *DNMT3A* CHIP^40^, the HRs of R882 mutations were higher than non-R882 mutations (i.e., non-R882 missense and pLoF together) for each (**Figure 2C**). R882 was associated with increased risk of persistent cytopenia, but not non-R882 mutations (R882, HR: 1.39, 95% CI: 1.11-1.74, P = 4×10^−3^ versus non-R882, HR: 1.09, 95% CI: 0.99-1.21, P=0.08) (**Figure 2D**). Similarly, only R882 was associated with increased risk of five-point MACE (R882, HR: 1.37, 95% CI: 1.20-1.56, P = 4×10^−6^ versus non-R882, HR: 1.07, 95% CI: 0.94-1.22, P=0.31) (**Figure 2D**). T Critically, these mutation-type-specific differences in clinical risk are not explained by differences in clone size. Median variant allele fractions did not significantly differ between TET2 pLoF and missense mutations (0.061 versus 0.059, P = 0.42) nor between DNMT3A R882 and non-R882 mutations (0.072 versus 0.062, P = 0.18), indicating that the observed heterogeneity in disease risk reflects intrinsic differences in enzymatic dysfunction rather than differences in clonal burden.

### Aberrant peripheral blood methylation of TET2 and DNMT3A CHIP associates with mutation type

Our data argue that mutations with the greatest impact on the enzymatic functions of *TET2* and *DNMT3A* confer the majority of disease risk; thus, we sought to examine whether DNA methylation itself from peripheral blood DNA might be a suitable readout of TET2 and DNMT3A enzymatic function and associated disease risk.

To identify sites across the genome that are differentially methylated in the setting of complete TET2 or DNMT3A loss, we introduced knock-out TET2 loss-of-function mutations, as in Kirmani et al^13^, and knock-in *DNMT3A* R882 mutations, as in Silver et al^41^, in CD34^+^ primary human hematopoietic stem and progenitor cells from mobilized peripheral blood of healthy donors (N = 6 for *DNMT3A*, N = 4 for *TET2*). After seven days in culture, these cells were flow sorted to isolate a purified population of CD34^+^CD38^−^Lin^−^ cells and assayed 5-methylcytosine profiles using duet evoC assay with the Twist Biosciences hybrid capture panel that targets ∼4 million CpG sites in the human genome^42^. We then performed an epigenome-wide association study (EWAS) for *TET2* loss-of-function and *DNMT3A* R882 mutations versus controls, respectively. Using regularized logistic regression, we then constructed two methylation-risk scores, one for *TET2* and one for *DNMT3A* (**Figure 3A**).

**Figure 3:**
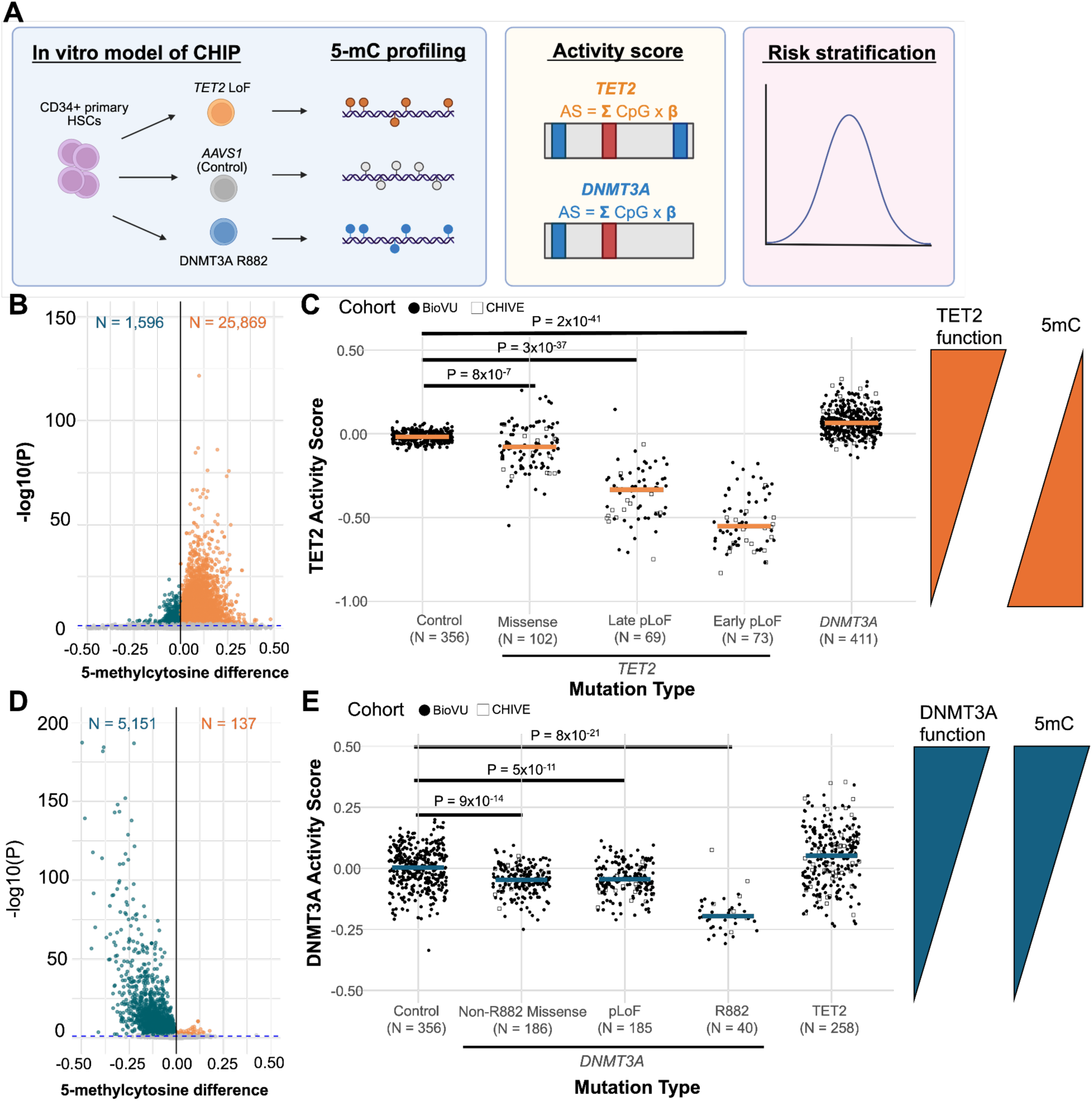
Derivation and validation of peripheral blood methylation-based activity scores. a) Experimental workflow: *AAVS1*-controlled CRISPR-Cas9 was used to generate *TET2* LoF and *DNMT3A* R882 models in primary human CD34^+^ HSCs (N=4–6), followed by 5-mC profiling to derive enzymatic activity scores (AS). b) Volcano plot of epigenome-wide association study (EWAS) results for *TET2* LoF, identifying 27,465 differentially methylated regions (DMRs; 500 bp). c) Validation of the TET2 activity score in the BioVU and CHIVE cohorts. The score shows a dose-dependent reduction corresponding to predicted pathogenicity: Early pLoF < Late pLoF < Missense < Control. d) EWAS results for *DNMT3A* R882 showing a predominance of hypomethylated regions (5,151/5,288). e) Validation of the DNMT3A activity score in patient cohorts. R882 variants exhibit the most profound enzymatic impairment. P-values determined by pairwise Wilcoxon rank-sum tests with Bonferroni correction.

For *TET2*, we identified 27,465 differentially methylated, 500 base-pair regions compared to controls. Expectedly, given TET2’s function as hydroxylating 5-methylcytosine, most were hypermethylated, with 25,869 regions differentially hypermethylated and 1,596 differentially hypomethylated (**Figure 3B**). Within the hypermethylated regions, we identified a subset of these CpGs profiled in common DNA methylation measurement arrays (e.g., Illumina Infinium Human Methylation-450 Beadchip 450 K array and Illumina EPIC array). Using peripheral blood DNA methylation profiles from the Illumina Infinium Human Methylation-450 Beadchip (450 K array) from the Framingham Heart Study (FHS), we used regularized regression to identify a set of 823 CpGs which predicted those with *TET2* pLoF (N = 74) versus controls (N = 148). Expectedly, a lower activity score in TET2 was associated with hypermethylation at the 823 sites.

To validate the relevance of the methylation-based metric, we applied the model to enzymatic methylation sequencing of peripheral blood DNA from two independent cohorts, Vanderbilt BioVU and the Clonal Hematopoiesis and Inflammation in the VasculaturE (CHIVE) cohort^43^ (BioVU baseline characteristics in **Supplemental Table 7**; CHIP mutations in the CHIVE cohort in **Supplemental Table 8**; distribution of variant allele fractions in **Extended Data Fig 3**). The resulting TET2 activity score exhibited a significant, dose-dependent decrease corresponding to the predicted pathogenicity of the *TET2* variants (**Figure 3C**). Notably, the degree of enzymatic impairment was stratified by the structural impact of the mutation: early pLoF mutations (proximal to amino acid residue 1129, preceding the catalytic domain) exhibited the most profound reduction in activity (N = 73, median TET2 activity score = -0.552), followed by late pLoF mutations occurring within or after the catalytic domain (N = 69, median TET2 activity score = -0.334). In contrast, missense mutations demonstrated a more attenuated effect on the score (N = 102, median TET2 activity score = -0.080), yet remained significantly distinct from non-CHIP controls (N = 356, median TET2 activity score = −0.019, P = 8×10^−7^). Pairwise comparisons confirmed that early pLoF variants were significantly more disruptive than both late pLoF (P = 2×10^−8^) and missense variants (P = 8×10^−27^), establishing the TET2 activity score as a high-resolution readout of variant-specific biochemical dysfunction. Interestingly, individuals with *DNMT3A* CHIP exhibited a significant elevation in TET2 activity score compared to controls (N = 411, median TET2 activity score = 0.063, P = 2×10^−71^).

For *DNMT3A* R882, we identified 5,288 differentially methylated, 500 base-pair regions compared to controls. Expectedly given DNMT3A’s function as a de novo methylation writer, most 5,151 regions were differentially hypomethylated and 137 differentially hypermethylated. Similar to the approach used for *TET2*, we developed a risk score for *DNMT3A* R882. Using peripheral blood DNA methylation profiles from the FHS, we fit a regularized regressionmodel. This model identified 542 CpGs that are predictive of individuals with *DNMT3A* R882 (N = 20) compared to controls (N = 40) among the CpGs profiled in common DNA methylation measurement arrays. These CpGs constitute the *DNMT3A* methylation risk score.

The *DNMT3A* R882-derived model, when applied to peripheral blood DNA from the BioVU and CHIVE cohorts (BioVU baseline characteristics in **Supplemental Table 7**; CHIP mutations in the CHIVE cohort in **Supplemental Table 8**; distribution of variant allele fractions in **Extended Data Fig 3**), revealed a DNMT3A Activity Score (DNMT3A AS) that stratified individuals by mutational archetype (**Figure 3E**). The most pronounced reduction in enzymatic activity was observed in carriers of the R882 hotspot mutation (N = 40, median DNMT3A activity score = -0.197), which was significantly lower than all other mutation classes (P < 2×10^−16^ for all pairwise comparisons). Notably, while both pLoF and non-R882 missense mutations exhibited significantly diminished scores relative to controls (N = 186, median DNMT3A activity score = -0.048 versus N = 356, median DNMT3A activity score = 0.002), these two groups were indistinguishable from each other in the DNMT3A activity score (P = 0.50). These data underscore the unique, potent dominant-negative effect of the R882 substitution on cellular methyltransferase activity compared to pLoF or other missense variants. Consistent with the reciprocal relationship observed in our TET2 analysis, *TET2*-mutated individuals demonstrated a significant elevation in DNMT3A activity score compared to controls (N = 258, median DNMT3A activity score = 0.051, P = 1.6×10^−6^). The finding that loss of TET2 function associates with a relative gain in the DNMT3A methylation signature, and vice-versa, suggests that these scores reflect the competing regulatory forces of de novo methylation and hydroxymethylation.

Importantly, while the TET2 and DNMT3A activity scores showed a modest but statistically significant correlation with variant allele fraction (TET2: Spearman rho = 0.21, P = 0.003; DNMT3A: Spearman rho = 0.17, P = 0.01), VAF explained less than 5% of the variance in either score, confirming that the methylation-based activity scores capture a dimension of enzymatic dysfunction that is largely orthogonal to clone size. In summary, this analysis demonstrated that *TET2* and *DNMT3A* mutation types observed to cause more disease clinically such as *TET2* pLoF and *DNMT3A* R882 also resulted in greater degrees of enzymatic impairment.

### Peripheral blood DNA methylation enzymatic activity is a personal biomarker of CHIP-associated disease risk

Given the observed heterogeneity in enzymatic impairment even within specific mutation classes, we hypothesized that an individual’s degree of *TET2* or *DNMT3A* activity could serve as a high-resolution, personal biomarker for CHIP-associated clinical outcomes. To test this, we evaluated the capacity of the enzymatic activity scores to predict incident persistent cytopenia and atherosclerotic cardiovascular disease within the BioVU cohort. Participants were stratified into high-risk (top 50%) and low-risk (bottom 50%) groups based on their methylation-derived activity scores, independent of their specific mutation type.

First, for incident persistent cytopenia, the five-year absolute risk was markedly higher among individuals with *TET2* CHIP and a high-risk TET2 activity score (15.9%, N = 17/107) compared to those with *TET2* CHIP and a low-risk TET2 activity score (6.6%, N = 7/106) and individuals without CHIP (3.4%, N = 12/356) (log-rank P < 0.001) (**Figure 4A**). The high-risk group was composed of 83% (89/107) TET2 pLoF mutations, whereas the low risk was composed of 50% (53/106) . In an adjusted Cox proportional hazards model, the high-risk TET2 activity score group was associated with a nearly five-fold increase in the risk of incident cytopenia compared to controls (HR = 4.99, 95% CI: 2.38-10.46, P = 2×10^−5^). Conversely, individuals carrying *TET2* mutations who exhibited a low-risk TET2 activity score showed an attenuated risk profile that did not significantly differ from the non-CHIP population (HR = 1.82, 95% CI: 0.68-4.85, P = 0.23).

**Figure 4:**
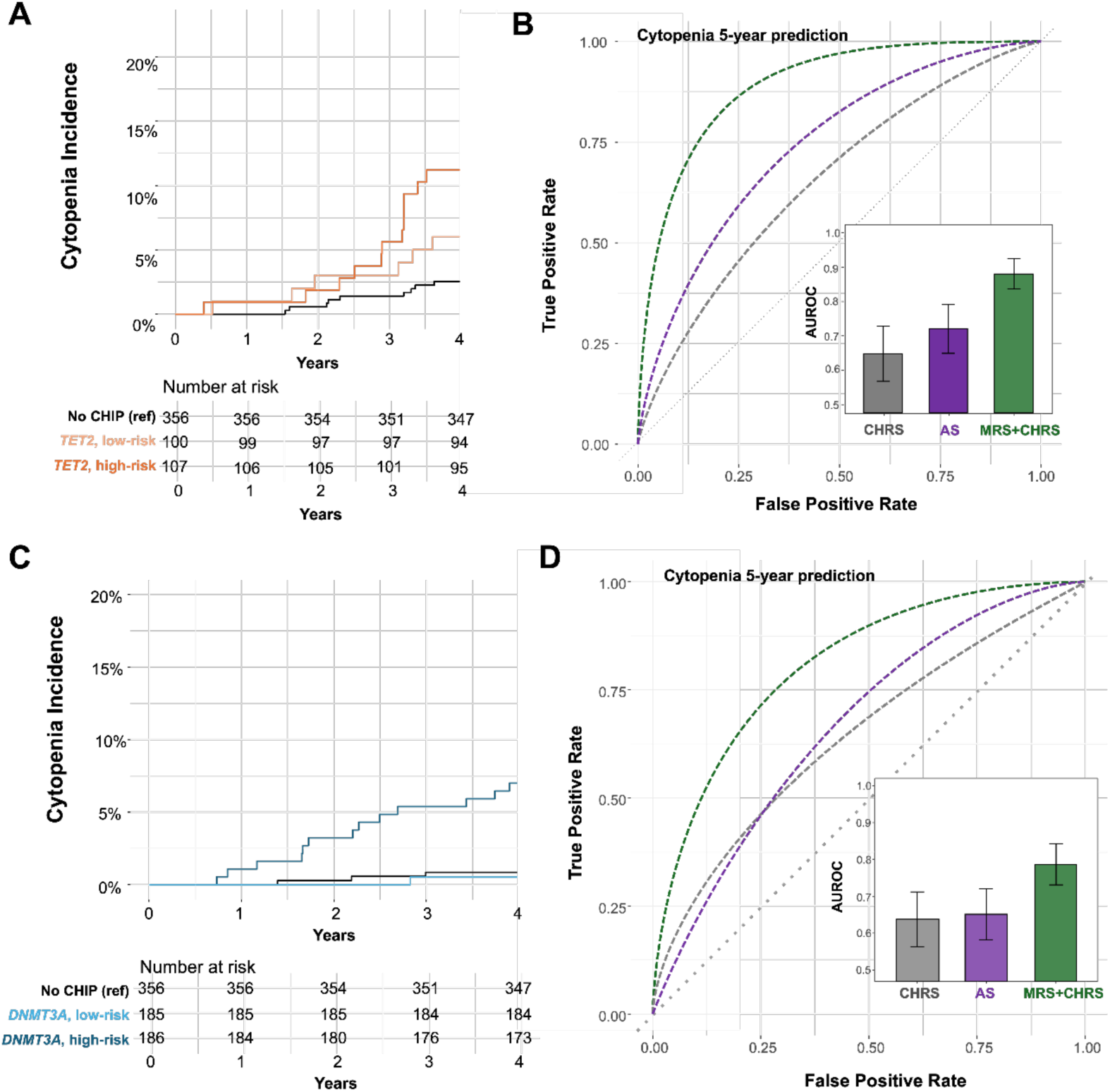
The enzymatic activity score as a functional predictor of incident cytopenia. a) Cumulative incidence of persistent cytopenia over 4 years in individuals without CHIP (N=356), *TET2* CHIP with low-risk activity score (AS) (N=106), and *TET2* CHIP with high-risk activity score (N=107; log-rank *p*<0.001). b) Receiver operating characteristic (ROC) curves comparing the predictive power of the Clonal Hematopoiesis Risk Score (CHRS) alone versus the combination of CHRS and TET2 AS. Inset: Area under the curve (AUROC) comparisons. c) Cumulative incidence of cytopenia stratified by DNMT3A activity score risk status (N=185–186 per CHIP group). d) ROC analysis demonstrating the synergistic predictive value of the DNMT3A activity score when added to the clinical CHRS model. Shaded areas represent 95% confidence intervals.

Notably, even among individuals with *TET2* pLoF mutations in the low-risk activity score group, the five-year cytopenia rate remained modestly elevated above the non-CHIP baseline (9.4%, 5/53 versus 3.4%, 12/356), For *TET2* missense mutations – which in aggregate were not associated with persistent cytopenia (**Figure 2B**) – the activity score identifies a subgroup of individuals at-risk: 11.1% (2/18) of those with *TET2* missense mutations in the high-risk group develop cytopenia at five years, compared to 3.8% (2/53) of those in the low-risk group. Together, these results demonstrate that the activity score refines rather than supplants mutation-specific information.

We next compared the predictive performance of the TET2 activity score against the Clonal Hematopoiesis Risk Score (CHRS), the current clinical standard for cytopenia and myeloid neoplasm risk. For the prediction of persistent cytopenia within five years, the CHRS achieved an area under the receiver operating characteristic curve (AUROC) of 0.655 (95% CI: 0.576–0.735). The TET2 activity score demonstrated marginally better independent performance (AUROC 0.728; 95% CI: 0.657–0.799). However, the integration of both metrics yielded a synergistic improvement in discrimination, with a combined AUROC of 0.885 (95% CI: 0.841 - 0.930, P<0.0001 vs. CHRS alone) (**Figure 4B**). These findings suggest that the methylation-based biomarker effectively captures the functional variance of *TET2* mutations and identifies those with *TET2* mutations who are at highest risk of disease from peripheral blood methylation.

Similarly, for *DNMT3A* CHIP and incident persistent cytopenia, the five-year absolute risk was markedly higher among individuals with *DNMT3A* CHIP and a high-risk DNMT3A activity score (16.7%, N = 31/186) compared to those with *DNMT3A* CHIP and a low-risk DNMT3A activity score (5.9%, N = 11/185) and individuals without CHIP (3.4%, N = 12/356) (log-rank P < 0.001) (**Figure 4C**). The high-risk group included 16% (30/186) *DNMT3A* R882 mutations, while the low-risk group included 5% (10/185) *DNMT3A* R882 mutations. Among R882 carriers concentrated in the high-risk group, the five-year cytopenia rate was 40.0% (12/30), compared to 20.0% (2/10) among R882 carriers in the low-risk group; this result suggest that the activity score provides additional resolution of individual disease risk even among those with R882 mutations. In an adjusted Cox proportional hazards model, the high-risk DNMT3A activity score group was associated with a nearly six-fold increase in the risk of incident cytopenia compared to controls (HR = 5.78, 95% CI: 2.90-11.5, P = 6×10^−7^). Conversely, individuals carrying *DNMT3A* mutations who exhibited a low-risk DNMT3A activity score showed an attenuated risk profile that did not significantly differ from the non-CHIP population (HR = 1.94, 95% CI: 0.84-4.48, P = 0.12). For *DNMT3A* non-R882 mutations, the activity score identifies a subgroup of individuals at-risk: 12.2% (19/156) of those with *DNMT3A* non-R882 mutations in the high-risk group develop cytopenia at five years, compared to 5.1% (9/175) of those in the low-risk group.

As for TET2, to evaluate the clinical utility of the DNMT3A AS, we compared its predictive power against the CHRS for persistent cytopenia within a five-year window. The standalone CHRS achieved an AUROC of 0.662 (95% CI: 0.593–0.731), while the DNMT3A activity score independently achieved an AUROC of 0.648 (95% CI: 0.574–0.723). However, the integration of the activity score with the CHRS significantly improved model discrimination, yielding a combined AUROC of 0.797 (95% CI: 0.742–0.853; P<0.0001 vs. CHRS alone) (**Figure 4D**).

Notably, while the CHRS already incorporates VAF as a continuous predictor, augmenting the CHRS with VAF alone provided no meaningful improvement in cytopenia discrimination for either *TET2* or *DNMT3A* CHIP (AUROC 0.658 versus 0.655, P = 0.71; and 0.665 versus 0.662, P = 0.68, respectively). These results indicate that the DNMT3A activity score captures critical functional heterogeneity not accounted for by clinical and demographic factors, providing a robust, synergistic biomarker for personalized risk assessment in DNMT3A-driven clonal hematopoiesis.

Second, for ASCVD, the TET2 activity score (TET2 AS) demonstrated a similar capacity to delineate clinical risk. Among individuals with TET2 CHIP, the high-risk TET2 activity score group exhibited a significantly elevated risk of incident ASCVD compared to controls (HR = 4.76; 95% CI: 2.25-10.06; P=4.4×10^−5^), while the low-risk group showed no significant increase in risk (HR = 1.18; 95% CI: 0.38-3.66; P=0.77) (**Figure 5A**). Similarly to cytopenia, among *TET2* missense mutations, the incidence of ASCVD at five years was 22.2% (4/18) in the high-risk group versus 11.3% (6/53) in the low-risk group. To determine the clinical utility of this functional readout, we compared the predictive performance of the TET2 activity score against the AHA PREVENT equations^10^, the current standard for cardiovascular risk assessment. In TET2 CHIP carriers, PREVENT alone achieved an AUROC of 0.723 (95% CI: 0.654-0.792), while the standalone activity score achieved an AUROC of 0.699 (95% CI: 0.626-0.773). Notably, the integration of the activity score with the PREVENT clinical model resulted in a substantial and significant improvement in risk stratification, yielding a combined AUROC of 0.930 (95% CI: 0.898-0.962; P < 1×10^−9^ vs. PREVENT alone) (**Figure 5B**). These results emphasize that the degree of enzymatic disruption, as captured by the TET2 AS, is a primary driver of the excess cardiovascular risk associated with *TET2* CHIP.

**Figure 5:**
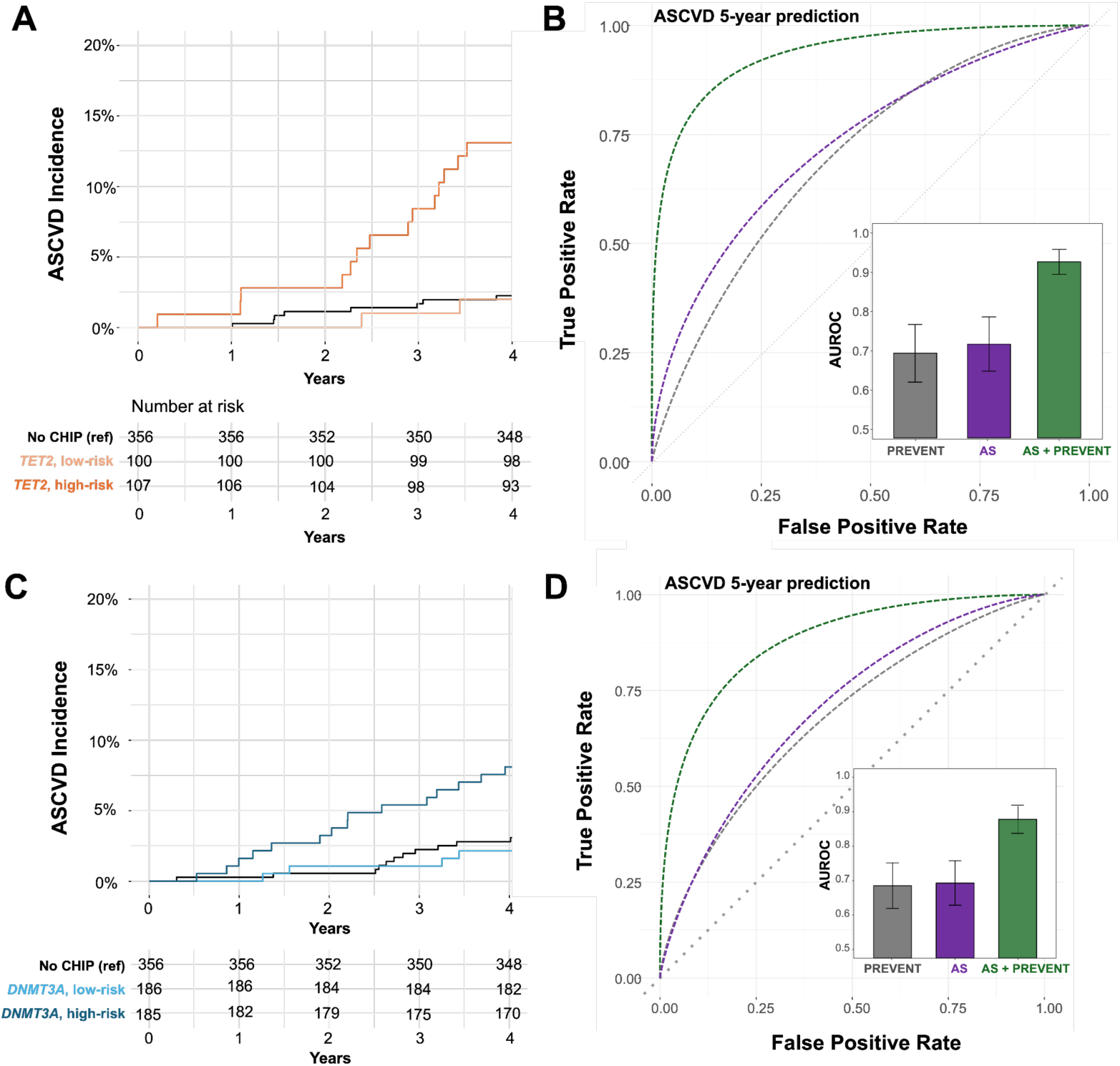
Functional methylation signatures significantly enhance cardiovascular risk stratification. a) Cumulative incidence of ASCVD in *TET2* CHIP carriers stratified by high-versus low-risk enzymatic activity. b) Comparison of the PREVENT versus the combined PREVENT and TET2 activity score model. The integrated functional model significantly outperforms standard clinical metrics (P < 1×10^−9^). c) ASCVD incidence by DNMT3A activity score risk status. d) ROC curves for *DNMT3A* CHIP carriers, illustrating the additive predictive value of the DNMT3A activity score over the PREVENT for incident cardiovascular events.

Finally, we assessed the relationship between the DNMT3A activity score (DNMT3A AS) and incident ASCVD. Within the *DNMT3A* CHIP cohort, individuals stratified into the high-risk DNMT3A activity score group exhibited an approximately two-fold increase in ASCVD risk compared to controls (HR = 1.99; 95% CI: 1.00-3.99; P=0.051), whereas those in the low-risk group showed no evidence of increased cardiovascular risk (HR = 0.71; 95% CI: 0.28-1.83; P=0.48) (**Figure 5C**). Similarly to cytopenia, among *DNMT3A* non-R882 mutations which do not associate with ASCVD (**Figure 2D**), the incidence of ASCVD at five years was 21.8% (34/156) in the high-risk group versus 8.6% (15/175) in the low-risk group. To evaluate whether this functional score provides additive value to established cardiovascular risk tools, we compared its performance against the PREVENT equation. In *DNMT3A* CHIP carriers, PREVENT achieved an AUROC of 0.695 (95% CI: 0.630-0.759), while the standalone DNMT3A activity score demonstrated similar discriminative power (AUROC 0.684; 95% CI: 0.618-0.750). Adding VAF to the PREVENT equation did not significantly improve ASCVD risk prediction (TET2: AUROC 0.731 versus 0.723, P = 0.38; DNMT3A: AUROC 0.703 versus 0.695, P = 0.44). Crucially, integrating the DNMT3A activity score with the PREVENT significantly enhanced the model’s predictive accuracy, yielding a combined AUROC of 0.875 (95% CI: 0.834-0.916; P < 1×10^−9^ vs. PREVENT alone) (**Figure 5D**).

## Discussion

Here, we leverage data from more than 1 million individuals across three diverse biobanks to demonstrate that residue-level mutational differences in TET2 and DNMT3A CHIP confer significantly different clinical risk profiles linked to their degree of enzymatic impairment. Surprisingly, we also find that an individual’s TET2 and DNMT3A enzymatic activity and thus clinical risk can be directly quantified from the methylation in peripheral blood. These observations yield three primary conclusions that redefine our understanding of disease risk attributable to clonal hematopoiesis.

First, the cellular and clinical consequences of *TET2* and *DNMT3A* CHIP are mutation-type-specific. We show that the pathogenic risk of TET2 CHIP across all disease outcomes is primarily concentrated in putative loss-of-function (pLoF) variants. Similarly, for DNMT3A, the dominant negative R882 mutation confers the majority of the disease risk. Furthermore, the selection pressures for acquiring loss of heterozygosity (CN-LOH) were mutation-specific, with TET2 pLoF variants exhibiting the strongest drive for homozygosity. Our finding that DNMT3A R882 mutations show no co-occurrence with CN-LOH provides the first molecular epidemiology evidence supporting its hypothesized dominant negative mechanism, consistent with structural studies showing that R882H preferentially forms stable homodimeric interfaces that prevent formation of functional wild-type/mutant heterotetramers^18,44^. These observations challenge current CHIP risk models that treat all mutations within a driver gene equally^7,8^, and underscore the necessity of mutation-level resolution for accurate risk prediction across the 75% of CHIP patients with mutations in *TET2* and *DNMT3A*. These data also provide a resolution to the ongoing debate regarding the association between CHIP and cardiovascular disease. Prior work has claimed that CHIP is not associated with increased risk of incident cardiovascular disease^45–47^, with some debate over CHIP detection methods^48^. However, by demonstrating that risk is concentrated in specific mutations, we show that the historical aggregation of all driver mutations may mask significant clinical associations and dilute the predictive power of *TET2* and DNMT3A CHIP as a cardiovascular biomarker.

Second, our findings suggest a biological threshold for pathogenicity that distinguishes clonal expansion from clinical disease. We conclude that the methylation-based activity scores capture a dimension of enzymatic dysfunction that is largely orthogonal to clone size.While many age-associated mutations in *TET2* and *DNMT3A* confer sufficient fitness for clonal expansion, only those exceeding a specific threshold of enzymatic impairment appear to drive distal clinical phenotypes (**Figure 6**). The degree of TET2 and DNMT3A enzymatic disruption provides a unifying functional mechanism for mutation-specific pathogenicity. Our data demonstrate that high-risk mutations (TET2 pLoF and DNMT3A R882) result in significantly greater enzymatic impairment, as quantified by our peripheral blood-based methylation risk scores, compared to lower-risk mutations. This directly links the mutational heterogeneity in clinical risk and cellular fitness to the magnitude of underlying enzymatic dysfunction.

**Figure 6:**
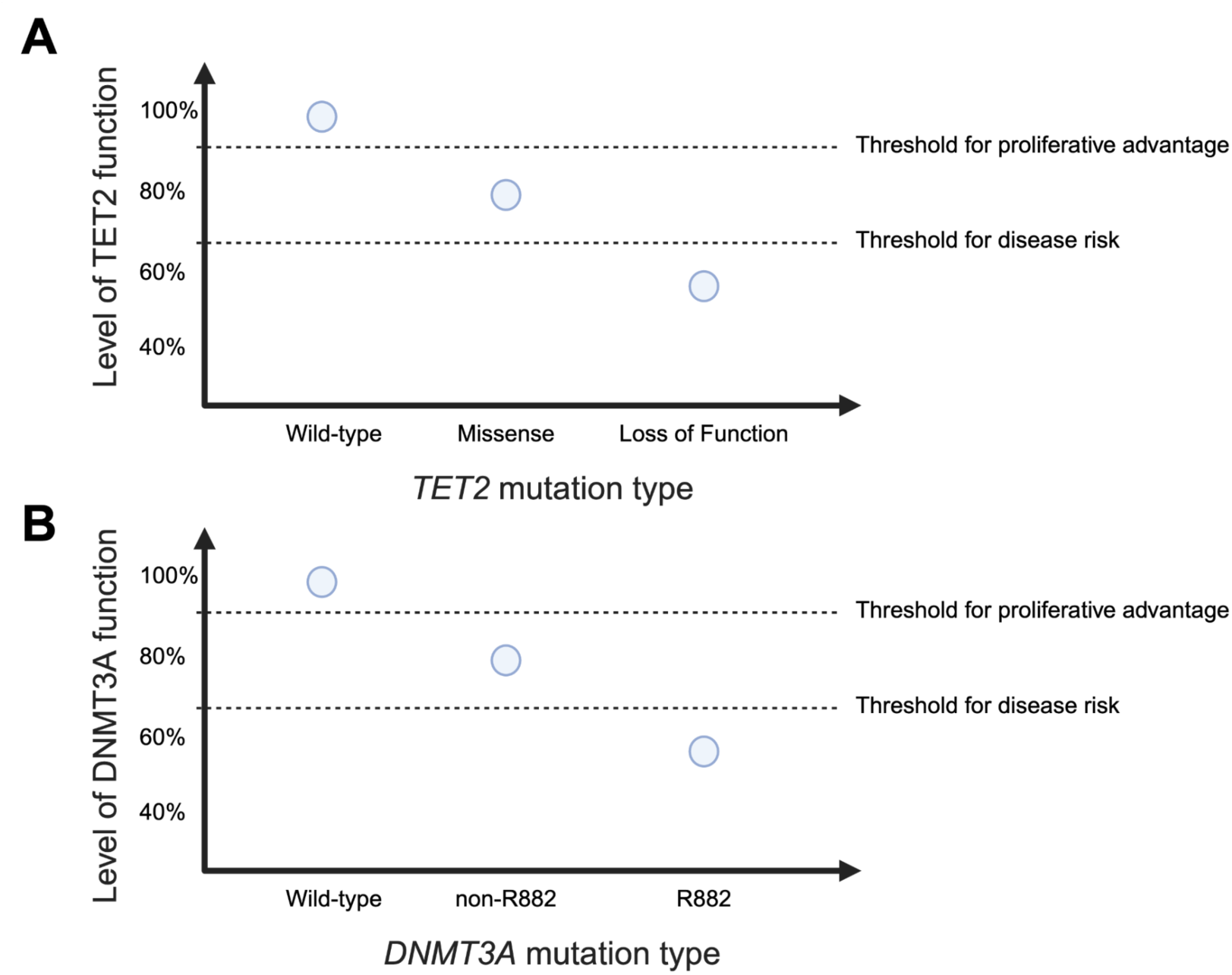
Proposed threshold model for clonal fitness and disease pathogenicity. Conceptual models for (a) *TET2* and (b) *DNMT3A*, illustrating the relationship between residual enzymatic function and clinical phenotype. A “threshold for proliferative advantage” allows for clonal expansion (CHIP), while a secondary, more stringent “threshold for disease risk” dictates the onset of distal clinical manifestations such as cytopenia and ASCVD. High-risk variants (pLoF, R882) uniquely cross this pathogenic threshold due to profound enzymatic disruption.

Third, an individual’s TET2 and DNMT3A enzymatic activity score acts as a superior, clinically useful personal biomarker to quantify CHIP-associated disease risk. We derived and validated an individual-level score that quantifies TET2 and DNMT3A activity from peripheral blood DNA methylation profiles. Incorporating this enzymatic activity score significantly outperformed existing clinical factor-based models in predicting ASCVD and incident cytopenia. Importantly, the synergy observed when combining our activity scores with established models indicates that functional epigenetic readouts capture a unique dimension of risk that clinical factors alone cannot resolve. Notably, while the CHRS already incorporates VAF as a binary predictor, augmenting the CHRS with VAF encoded continuously provided no meaningful improvement in cytopenia discrimination for either *TET2* or *DNMT3A* CHIP. Similarly, adding VAF to the PREVENT equation did not significantly improve ASCVD risk prediction. In contrast, the activity score improved combined model performance for cytopenia and ASCVD; these improvements in predictive value are not attributable to clone size and instead reflect the independent prognostic value of enzymatic dysfunction quantified from peripheral blood methylation. The methylation activity scores therefore represent a new class of biomarker for stratifying CHIP patients for prognosticating risk and prospectively testing therapeutic interventions.

This study has several limitations. The reliance on retrospective biobank data, which, on one hand, provides significant statistical power, may introduce survival bias, has heterogeneity in endpoint assessment, and limits sensitivity for detection of small CHIP clones^49^. Further, the population in these biobanks may not be broadly representative. Additionally, the specific mechanisms by which the observed DNA methylation changes lead to distal phenotypes like ASCVD require further investigation. Future work should prospectively validate these activity scores in clinical trial settings. Finally, CHIP detection in the BioVU cohort was derived from whole-genome sequencing at approximately 30x mean depth, which limits sensitivity to clones with variant allele fractions below 0.02 and enriches the analytic cohort for larger, more established clones relative to what would be detected by error-corrected targeted sequencing^49^. The clinical associations observed here may therefore not fully reflect the risk profile of early, small CHIP clones detectable only by more sensitive sequencing approaches

In conclusion, our study of over 1 million individuals demonstrates that the clinical risk from TET2 and DNMT3A clonal hematopoiesis is highly dependent on the specific mutation type. This mutation-specific pathogenicity is mechanistically linked to the magnitude of underlying enzymatic disruption, which can be quantified by a peripheral blood DNA methylation based TET2 and DNMT3A activity score. This enzymatic activity score represents a novel and superior personal biomarker to accurately stratify an individual’s CHIP-associated disease risk and may be broadly useful for quantifying the pathogenicity of other genes involved in DNA methylation.

## Methods

### Cohorts for mutation-specific epidemiology analyses

This study utilized data from three large-scale cohorts: Vanderbilt BioVU, NIH AllofUs (AoU) Research Program, and the UK Biobank (UKB). Participants with available whole genome or exome sequencing data were initially considered. We excluded individuals with a history of hematologic malignancy or other non-neoplastic clonal disorders (**Supplementary Table 9**), lack of follow-up information, or missing key covariates. As CHIP is highly uncommon in younger individuals, to minimize potential artifacts, we excluded participants with baseline age <40. Our final analytic cohort was 846,353 participants across BioVU, AoU, and UKB (**Extended Data Figure 1**).

### Clonal hematopoiesis of indeterminate potential detection

Putative somatic SNVs and short indels were called with GATK Mutect2 and filtered according to previously established criteria among BioVU, AoU, and UKB, as we have previously described^5^. Briefly, 74 canonical CHIP genes were screened for potential CHIP mutations using the Mutect2 somatic variant caller as described in Vlasschaert et al (**Supplemental Table 10**).^2^ Variants included in the preliminary dataset met the following criteria: presence in a pre-established list of candidate CHIP variants, total sequencing depth ≥ 20, alternate allele read depth count ≥ 5, and representation in both sequencing directions (i.e., F1R2 ≥ 1 and F2R1 ≥ 1). CHIP mutations were defined as those with a variant allele fraction (VAF) ≥ 0.02. CHIP detection across all cohorts was derived from whole genome/exome sequencing data, using a consistent detection method and the same canonical CHIP driver genes list, which specifies candidate missense and indel variants for each gene, as well as a set of genes in which truncating and splice site variants may be considered.

### Definition of mutation types in TET2 and DNMT3A

Using the annotations from ANNOVAR, mutation-specific analyses were performed for variants in *TET2* and *DNMT3A*. Observed mutations were categorized into distinct archetypes based on their predicted structural and functional impact. *TET2* variants were stratified into two primary groups: (1) putative loss-of-function mutations (pLoF), including all nonsense (stop-gain) mutations and frameshift insertions or deletions. pLoF variants were further subdivided into early pLoF (proximal to amino acid residue 1129, preceding the catalytic domain) and late pLoF (occurring within or distal to the catalytic domain) and (2) missense mutations, including all non-synonymous missense mutations identified within the canonical *TET2* coding sequence that met the CHIP inclusion criteria^23,24,26^. Given the high prevalence and unique dominant-negative biochemical properties of the R882 hotspot, *DNMT3A* variants were stratified into two groups: (1) R882 mutations, specifically including the R882H and R882C missense substitutions, and (2) non-R882 mutations. This aggregate group included all other *DNMT3A* mutations qualifying as CHIP, including non-R882 missense mutations, splicing variants, and protein-truncating variants (frameshift and stop-gain)^5,11,16,18^.

### cBioPortal myeloid neoplasm analysis

To compare the mutational landscape of CHIP with overt myeloid neoplasm, we aggregated mutational data from 16,264 cases of myeloid neoplasms (MN) available via the cBioPortal for Cancer Genomics. We extracted protein-level coordinates for all TET2 and DNMT3A mutations. Differences in the distribution of mutation types (pLoF vs. missense) and specific residue hotspots between CHIP and MN cohorts were assessed using Fisher’s exact tests with Bonferroni correction for multiple hypothesis testing.

### Copy-neutral loss of heterozygosity detection and analysis

As described in Zhao et al, autosomal mosaic chromosomal alterations (mCAs) were detected using MoChA. In the UKB, mCAs were detected from genotyping array data from the same DNA samples used for CHIP detection, as previously described by Loh et al^50,51^. Similarly, in BioVU, the Illumina Expanded Multi-Ethnic Genotyping Array (MEGAEX) were used to call mCAs, as described by Kishtagari et al^28^. In All of Us, for the version 8 of the genotyping data, detection of mCAs was performed using the Infinium Global Diversity Array with the same methods as performed in UKB and BioVU. MoChA utilizes genotypes to evaluate coverage and BAF at heterozygous loci, identifying mCAs based on deviations in allelic balance. MoChA was run with the additional parameter –LRR-weight 0.0 –bdev-LRR-BAF 6.0, deactivating the LRR + BAF model to improve detection sensitivity.^52^ Cellular fitness was inferred by calculating the odds ratio of a specific mutation type co-occurring with CN-LOH on the same chromosomal arm (i.e., *DNMT3A* and 2p and *TET2* and chr4q) compared to the background rate of CN-LOH in individuals without CHIP.

### Phenome-wide association studies and meta-analysis

We conducted time-to-event phenome-wide association studies (PheWAS) in BioVU, AoU, and UKB to evaluate associations between specific mutation types in *DNMT3A* and *TET2* and a comprehensive range of clinical outcomes. Phenotypes were defined using Phecodes (https://phewascatalog.org/phewas/#phex) mapped from International Classification of Diseases, Ninth and Tenth Revision (ICD-9/ICD-10) codes curated from electronic health records (EHRs). For each individual, the presence of any ICD codes corresponding to a given Phecode inclusion criterion classified the subject as a case; absence of such codes classified the individual as a control. Individuals with prevalent diagnoses prior to DNA collection were excluded from all time-to-event disease-association analyses.

We performed a time-to-event PheWAS across 2,180 clinical phenotype. That is, we fitted Cox proportional hazards models (R package “survival”) to estimate HRs and 95% CIs for the association between mutations in TET2 or DNMT3A (i.e., pLoF/missense and R882/non-R882) and incident disease risk. Models were adjusted for age at blood draw (continuous), age squared (continuous), genetic sex (categorical), current smoking status (categorical), and principal components 1–10 (continuous). PheWAS analyses were performed separately in BioVU, AoU, and UKB and subsequently combined using inverse-variance weighted fixed-effects meta-analysis, as implemented in the R package “metafor.” All P values were corrected for multiple testing using the Bonferroni method. For the comparison of mutation-specific effect sizes, we employed a mixed-effects meta-regression to account for heterogeneity across phenotypes.

### Specific outcome analysis by mutation type

As initially defined by Brogan et al^4^, persistent cytopenia was defined as having two consecutive abnormal blood counts in a single cell lineage occurring at least 120 days apart, without any normal measurements in between. Following modified World Health Organization (WHO) criteria, thresholds for abnormal counts were set at hemoglobin <12.0 g/dL (females) or <13.0 g/dL (males) for anemia, platelets <150,000 cells/mL for thrombocytopenia, and white blood cell count <3700 cells/mL for leukopenia. The date of onset was recorded as the first occurrence of the persistent cytopenia. To analyze risk, we fitted Cox proportional hazards time-to-event models to estimate HRs and 95% CIs for the association between mutations in *TET2* or *DNMT3A* (i.e., pLoF/missense and R882/non-R882) and incident persistent cytopenia risk. Similarly, we defined five-point major adverse cardiovascular events (MACE) as the composite of coronary artery disease, heart failure, stroke, peripheral vascular disease or death.

### In vitro model of TET2 loss-of-function and DNMT3A R882 clonal hematopoiesis

As described in Kirmani et al^13^ and Silver et al^41^, we utilized primary human CD34^+^ hematopoietic stem and progenitor cells (HSPCs) derived from mobilized peripheral blood of healthy donors (Fred Hutchinson Cancer Research Center; StemCell Technologies). Cells were expanded for 48 hours in StemSpan II medium supplemented with CD34+ expansion supplement, UM729 (500 nM), and Stemreginin-1 (750 nM) prior to editing.

We introduced *TET2* mutations via CRISPR-Cas9 ribonucleoprotein (RNP) electroporation. For *DNMT3A* R882, two homology donor (HD) templates with distinct self-cleaving fluorescent tags were used and fluorescence-activated cell sorting (FACS) was used to isolate homozygous R882H- or R882C-mutant cells. RNP complexes were formed by incubating Alt-R HiFi spCas9 Nuclease V3 with gene-specific sgRNAs (IDT) at a 1:3.26 ratio. Electroporation was performed using a Neon Transfection System (1650 V, 10 ms, 3 pulses). Editing efficiency was confirmed by Sanger sequencing and TIDE analysis of genomic DNA. Seven days post-editing, the primitive CD34^+^CD38^−^Lin^−^ cell population was isolated via FACS.

To profile 5-methylcytosine, we extracted genomic DNA from sorted cells and generated duet evoC libraries (biomodal) for five-letter next-generation sequencing (A, C, G, T, methyl-C). Libraries were enriched for CpG sites using the Twist Human Methylome Panel and sequenced on an Illumina NovaSeq 6000 (150 bp paired-end). Raw data were processed through the modality biomodal pipeline (v1.1.1) to quantify the modification state of each CpG site across the genome (GRCh38).

### Epigenome-wide association study

To identify differentially methylated regions (DMRs), we performed an epigenome-wide association study using the modality framework, which analyzes DNA modification data by modeling the underlying probability distributions of methylation states. We segmented the genome into 500 base-pair (bp) non-overlapping windows. For each 500bp window, we aggregated the modification probabilities of all CpG sites within that range to generate a regional methylation profile. We then compared the methylation distributions between the mutant groups (*TET2* loss-of-function or *DNMT3A* R882) and the non-edited controls. To identify a consensus signature for *DNMT3A* R882, we integrated results from both R882C and R882H variants, we calculated the average 5-methylcytosine (5mC) difference and the mean P-value across the R882C and R882H models. Regions were defined as DMRs if they reached a significance threshold of Benjamini-Hochberg adjusted P < 0.05 in both R882C and R882H for *DNMT3A* and in *TET2* loss-of-function.. The modality tool was used to calculate a divergence metric (Jensen-Shannon Divergence) to quantify the difference between these distributions. Regions were defined as DMRs if they had a Benjamini-Hochberg adjusted P-value < 0.05).

### Enzymatic activity score construction

To quantify individual-level enzymatic dysfunction from peripheral blood DNA methylation, we constructed TET2 and DNMT3A Activity Scores (AS) using a multi-step regularized regression pipeline. We first restricted our analysis to the CpG sites contained within the 500 bp differentially methylated regions (DMRs) identified in our in vitro models (27,465 regions for TET2; 5,288 regions for DNMT3A). To ensure portability with clinical datasets, we further filtered for CpGs profiled on standard DNA methylation arrays (Illumina HumanMethylation450 and EPIC). To minimize redundancy and handle multicollinearity, we performed correlation-based pruning. DMRs were ranked by their association significance, and any neighboring region with a correlation coefficient R^2^ > 0.8 was excluded, retaining only the most statistically robust representative from the block of associated CpGs.

The Activity Scores were trained using peripheral blood DNA methylation profiles from the Framingham Heart Study (FHS) as a discovery cohort. We utilized Elastic Net regularized logistic regression (L1/L2 ratio = 0.5) to identify the optimal parsimonious set of methylation features that predicted mutational status. The TET2 activity score was trained to discriminate participants with TET2 pLoF (N=74) from age- and sex-matched controls (N=148), resulting in a final model comprising 823 weighted CpGs. The DNMT3A activity score was trained to discriminate participants with DNMT3A R882 mutations (N=20) from matched controls (N=40), resulting in a final model of 542 weighted CpGs.

Individual activity scores were calculated by computing the dot product of the standardized methylation beta-values (X_scaled_) and the non-zero model coefficients (weights):

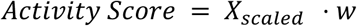

The resulting continuous scores provide a high-resolution readout of biochemical dysfunction. For all downstream clinical analyses, the scores were standardized to the non-CHIP control population (mean = 0, SD = 1).

*Validation in Vanderbilt BioVU cohort and Clonal Hematopoiesis and Inflammation in the VasculaturE (CHIVE) cohorts* We then applied our enzymatic activity scores to participants from BioVU, Vanderbilt’s biorepository of DNA linked to de-identified electronic health records, and participants from the Clonal Hematopoiesis and Inflammation in the VasculaturE (CHIVE) cohort consisted of patients recruited at Vanderbilt University Medical Center^43^ to validate variant-specific biochemical dysfunction. We obtained cellular DNA from peripheral whole blood in BioVU from 356 individuals who did not have CHIP, 207 individuals with *TET2* CHIP, and 371 individuals with *DNMT3A* CHIP (**Supplemental Table 7**) and in CHIVE from 50 individuals with *TET2* CHIP and 40 individuals with *DNMT3A* CHIP (**Supplemental Table 8**).

DNA methylation was profiled using a high-throughput, cost-effective targeted enzymatic methyl-seq (EM-seq) assay via NEBNext Enzymatic Methyl-seq kit paired with a Twist Bioscience hybrid capture panel targeting approximately 4 million functionally relevant CpG sites^42^. Libraries were sequenced on the Illumina NovaSeq 6000 platform to a mean depth of 30x. Reads were aligned to the human reference genome (GRCh38). Beta matrices were generated from raw CX report files using an optimized WDL-based bioinformatic pipeline. Among CpG sites meeting a minimum coverage threshold of 10x in each sample, we selected CpG sites where > 80% of the cohort had ≥ 25x coverage. Individual beta values, represented as the ratio of methylated reads to total coverage, were then extracted and merged into a single beta standardized matrix. After subsetting the beta matrices to the CpG sites in the activity scores, we computed TET2 and DNMT3A activity scores for the individuals in BioVU and CHIVE and compared them by mutation type with pairwise Wilcoxon rank-sum tests.

### Clonal hematopoiesis risk score calculation

Prior work by Weeks et al^7^ has described the Clonal Hematopoiesis Risk Score (CHRS), which stratifies risk of CHIP transformation into leukemia and all-cause mortality in the UK Biobank. The CHRS uses features such as mutation count, driver gene, VAF, age, cytopenia, and cell morphology to prognosticate risk. Participants were called low-risk for CHRS ≤ 9.5, intermediate-risk for 10 ≤ CHRS ≤ 12, and high-risk CHRS ≥ 12.5.

### PREVENT equation calculation

For cardiovascular risk assessment, we calculated the risk of atherosclerotic cardiovascular disease (ASCVD) using the AHA PREVENT (Predicting Risk of cardiovascular Disease events) equations. Due to limited follow-up time, we applied the PREVENT risk score to predict 5-year risk. The PREVENT equations were selected as they represent the updated standard for cardiovascular risk estimation, notably removing race as a predictor while incorporating measures of renal function. Variables included in the calculation were age, sex, systolic blood pressure, blood pressure treatment status, current smoking status, diabetes, total cholesterol, and HDL cholesterol. Additionally, where available, estimated glomerular filtration rate (eGFR) was included to enhance risk prediction. Scores were calculated using clinical data from the encounter nearest to the date of genetic sequencing.

### Time-to-event analyses

We utilized Cox proportional hazards models to estimate hazard ratios (HR) for the association between CHIP mutation types, enzymatic activity scores, and incident clinical outcomes (ASCVD, persistent cytopenia, and five-point MACE). All models were adjusted for age, sex, smoking status, and the first ten principal components of ancestry. ASCVD models were additionally adjusted for BMI, LDL-C, diabetes, hypertension, and statin use. Follow-up time was defined from the date of sequencing until the first event occurrence, death, or the date of the last medical record update.

### Evaluation of predictive risk models

The incremental value of the enzymatic activity scores (AS) was evaluated by comparing the performance of clinical-only models (CHRS for cytopenia; PREVENT for ASCVD) against integrated models (Clinical + AS). Model performance was quantified using the Area Under the Receiver Operating Characteristic curve (AUROC) for 5-year incident risk.

## Data Availability

Individual-level sequence data and CHIP calls have been deposited with UK Biobank and are freely available to approved researchers, as done with other genetic datasets to date under data fields (30105, 30106, 30107). The genotypes and phenotypes of UKB and AoU participants are available by application to the UKB (https://www.ukbiobank.ac.uk/register-apply/) and AoU (https://allofus.nih.gov/), respectively. For Vanderbilt BioVU, clonal hematopoiesis of indeterminate potential sequencing calls are available through controlled access to qualified researchers. Due to Vanderbilt BioVU cohort restrictions, access will require a data use agreement with Vanderbilt University Medical Center, which can be facilitated by the corresponding author, Dr. Bick. Methylation data for BioVU is available via dbGaP at accession number phs004433.v1.p1. CHIP calls and methylation data for CHIVE is available upon request. Framingham Heart Study methylation data is available publicly for researchers at dbGaP accession number phs000974.v1.p1.

## Code Availability

Code for the study is available at https://github.com/bicklab/mut-specific-methylation.

## Acknowledgements

This work was supported by the following National Institutes of Health grants: UG3 AG097155 (A.G.B., M.R.S.), K08 HL171833 (J.B.H.), R01 AG088657 (A.G.B.), F30 AG099331 (Y.P.), and T32 GM145734 (J.C.V.A.). Additionally, this work was supported by a Burroughs Wellcome Fund Career Award for Medical Scientists (A.G.B.), a Pew-Stewart Scholar for Cancer Research award (A.G.B.), supported by the Pew Charitable Trusts and the Alexander and Margaret Stewart Trust (A.G.B.), a Hevolution/AFAR New Investigator Award in Aging Biology and Geroscience Research (A.G.B.), and Arthritis National Research Foundation grant 1288083 (R.W.C.). The sequencing of 250,000 WGS individuals from BioVU® has been funded by the Alliance for Genomic Discovery consisting of NashBio, Illumina and industry partners Amgen, AbbVie, AstraZeneca, Bayer, BMS, GSK, Merck, and Novo.

## Competing Interests

M.R.S. has received honoraria from Bristol Myers Squibb, CTI, Forma, Geron, GlaxoSmithKline, Karyopharm, Ryvu, and Taiho; research funding from ALXOncology, Astex, Incyte, Takeda, and TG Therapeutics; holds equity in Empath Biosciences, Karyopharm, and Ryvu; and travel reimbursement from Astex. All other authors declare no competing interests.

## Author Contributions

Y.P. and K.Z. contributed equally to this work. Y.P. and K.Z. performed data analysis. J.C.V.A, A.J.S., and Y.A. performed experiments. A.K., M.R.S., R.W.C., J.B.H., and A.G.B. contributed clinical samples. R.W.C., E.H., J.B.H., and A.G.B. supervised epigenetic analyses. J.B.H. and A.G.B. jointly supervised this work.

## Extended Data Figures

**Extended Data Figure 1:**
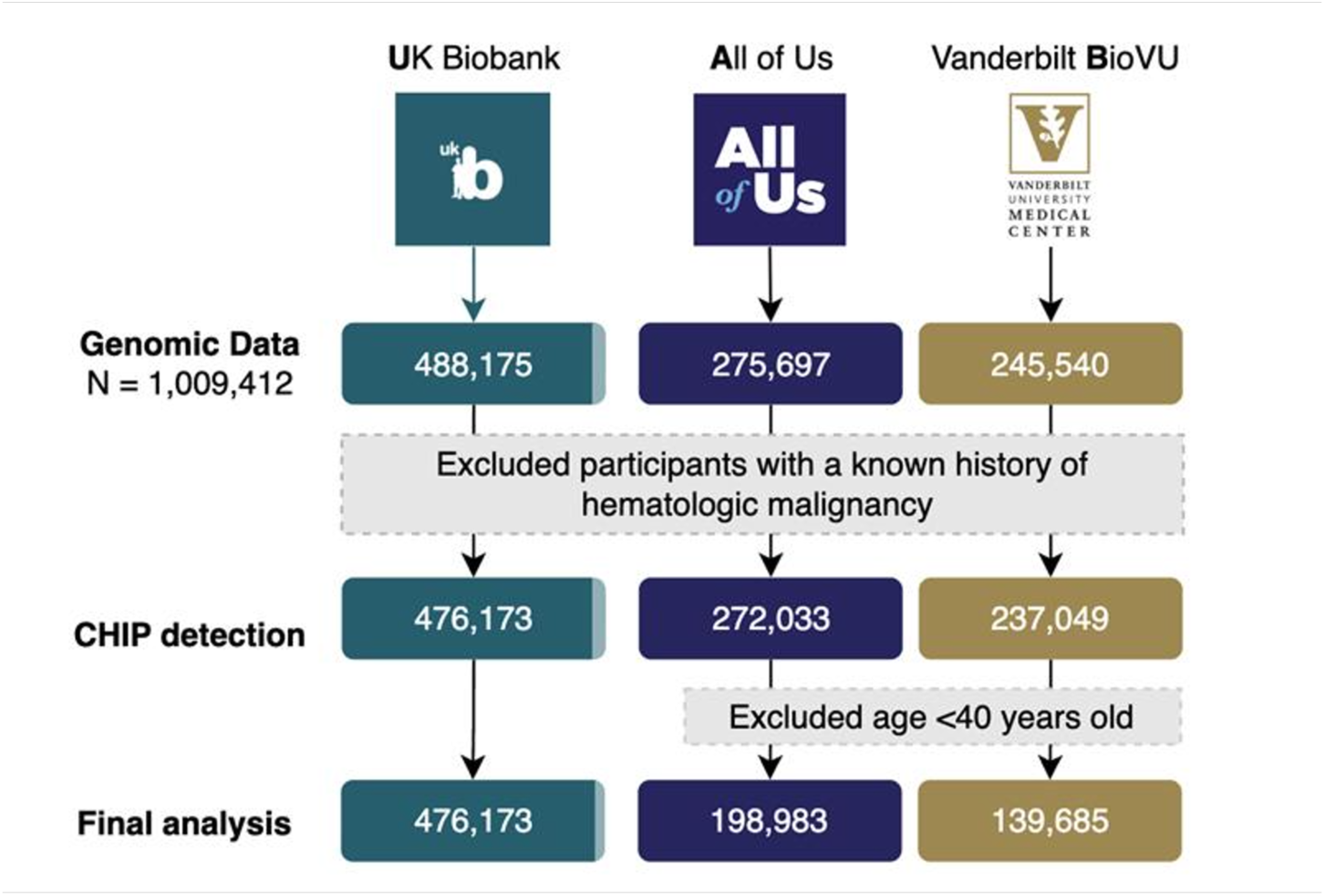
Flow diagram showing participant selection in the All of Us Research Program, Vanderbilt BioVU, and UK Biobank cohorts. Individuals were excluded if they had (1) a history of hematologic malignancy or other non-neoplastic clonal disease, (2) missing follow-up information, or (3) missing key covariates. Participants younger than 40 years were further excluded in All of Us and BioVU.

**Extended Data Figure 2:**
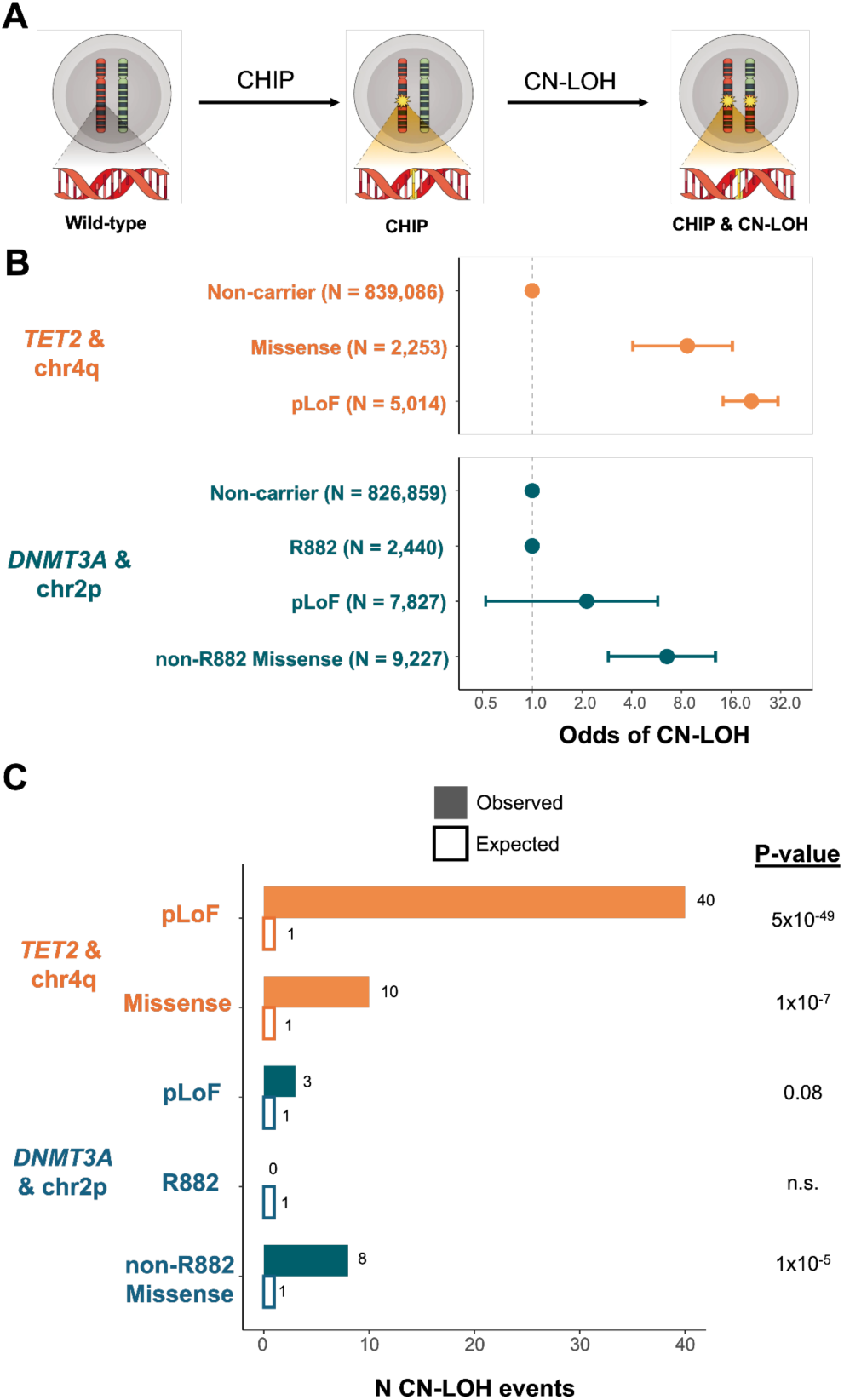
Mutation-specific selection pressure and drive for copy-neutral loss of heterozygosity (CN-LOH). b) Schematic representing the acquisition of somatic CHIP mutations followed by a second-hit CN-LOH event, leading to cellular homozygosity. c) Forest plots showing odds ratios (OR) for the co-occurrence of CN-LOH at chr4q (*TET2*) and chr2p (*DNMT3A*) across mutational subtypes. Error bars represent 95% confidence intervals. d) Observed versus expected frequency of CN-LOH events. *TET2* pLoF variants show the strongest drive for selection (P=5×10−49), whereas *DNMT3A* R882 mutations exhibit zero co-occurrence with chr2p CN-LOH. This lack of selection pressure for homozygosity supports the hypothesized dominant-negative function of the R882 substitution. P-values determined by two-sided Poisson test.

**Extended Data Figure 3:**
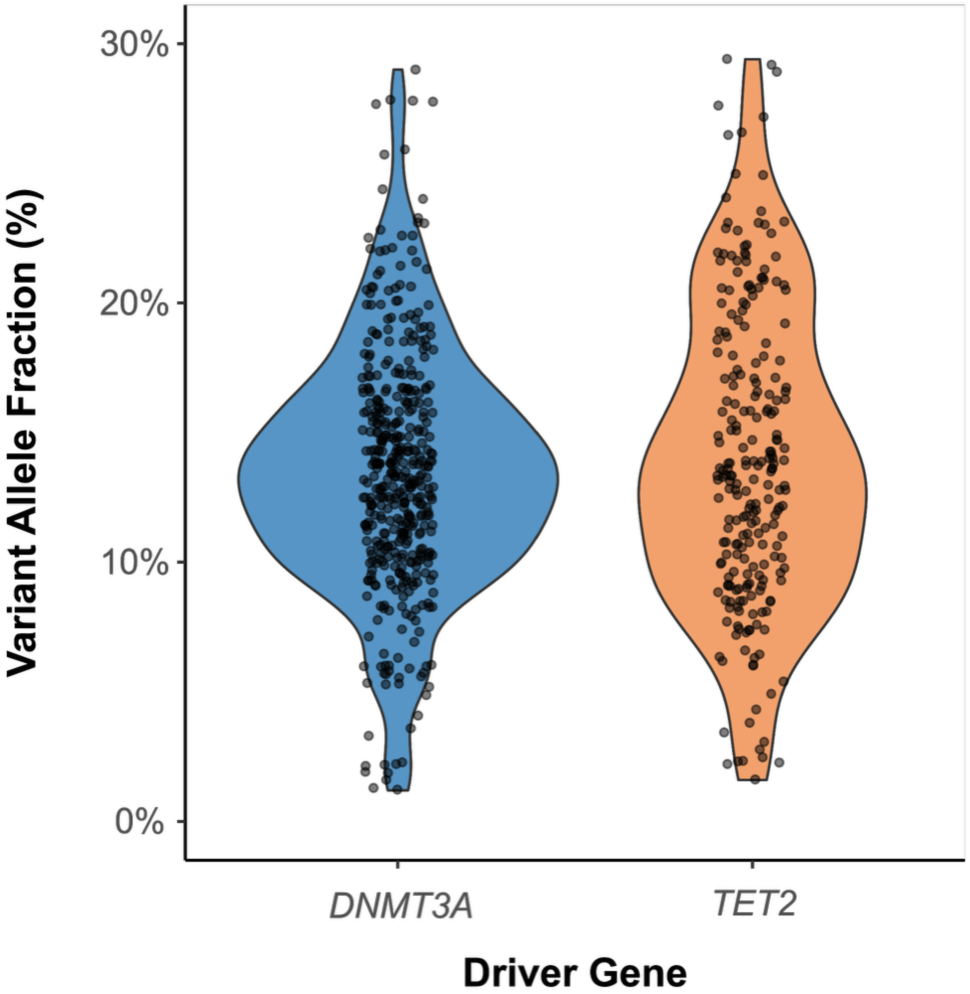
Distribution of variant allele fractions (VAFs) stratified by driver gene (*DNMT3A* and *TET2*) for participants in the BioVU and CHIVE studies.

## Notes

### Funding Statement

This study was funded by the following National Institutes of Health grants: UG3 AG097155 (A.G.B., M.R.S.), K08 HL171833 (J.B.H.), R01 AG088657 (A.G.B.), F30 AG099331 (Y.P.), and T32 GM145734 (J.C.V.A.). Additionally, this work was supported by a Burroughs Wellcome Fund Career Award for Medical Scientists (A.G.B.), a Pew-Stewart Scholar for Cancer Research award (A.G.B.), supported by the Pew Charitable Trusts and the Alexander and Margaret Stewart Trust (A.G.B.), a Hevolution/AFAR New Investigator Award in Aging Biology and Geroscience Research (A.G.B.), and Arthritis National Research Foundation grant 1288083 (R.W.C.). The sequencing of 250,000 WGS individuals from BioVU has been funded by the Alliance for Genomic Discovery consisting of NashBio, Illumina and industry partners Amgen, AbbVie, AstraZeneca, Bayer, BMS, GSK, Merck, and Novo.

